# The Impact of Social Distancing on The Course of The Covid-19 Pandemic in Four European Countries

**DOI:** 10.1101/2020.05.03.20089680

**Authors:** Andrew Shardlow

## Abstract

In this article the mortality data from four European countries arising from the Covid-19 pandemic is modelled using logistic functions. The countries chosen for examination are Spain, Italy, France and the UK. They have been selected because in each the pandemic is advanced, mortality high and any prospect of containment has passed. They have also been selected because in each social distancing has been used in an attempt to reduce peak daily mortality with relatively strict enforcement following a defined date. The choices of data set and model type is justified. The impact, if any, of social distancing is examined.

## 2 The Selection of a Covid-19 Pandemic Model

It is probable Covid-19 infection has a high proportion of asymptomatic or minimally symptomatic[1][2] persons and is infectious before symptom development.[3] This has allowed the disease to spread widely amongst the population in countries where early containment was not successful or not attempted. In these circumstances it is reasonable to use a simple pandemic model to provide an overview of pandemic progress. This is in contradistinction to the complex numerical modelling with geographic inputs and outputs that provide detailed local morbidity data to inform logistics and planning. Mechanistic epidemiological models are often founded on unknown or imprecisely known parameters and in this sense are not precisely dynamic. They do however inform the mathematical form of kinetic models.

### 2.1 The Kermack and McKendrick Model

As this disease is of short duration in relation to population life expectancy we might use the *SIR* compartmental model of Kermack and McKendrick.[4] This is a three compartment model in which *N* represents the size of the population of interest, *S* represents the number of susceptible people, *I* those on their infective journey to *R*, the recovered or dead. In the following equations *β* represents the product of the average contact rate *c* and *p_i_* the probability of transmitting infection in a single contact between an infected and a susceptible person. *γ* is the rate of transition from *I* to *R*. It is defined by three ordinary differential equations:

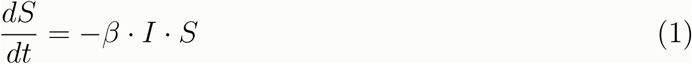

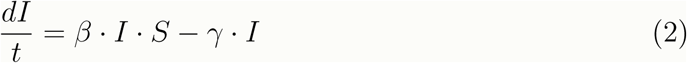

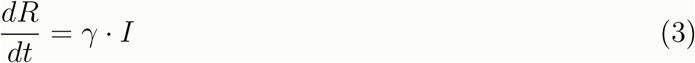

On reflection however, the assumption present in equation 3 is only valid if illness duration is uniformly distributed within *I*. This is only true if *I* is constant over time, which in general is not the case.

### 2.2 A Modified Kermack and McKendrick Model

An alternative *SIR* model can be developed in which *R* is a time delayed convolution of *N* − *S* with a normalized mask *M* with its peak shifted forward in time by *λ*, the latency in *I*, and representing the probability over time of leaving *I* after entry into *I*.

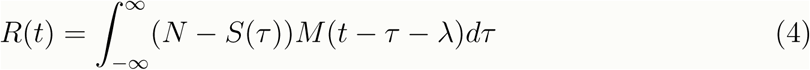

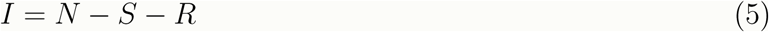

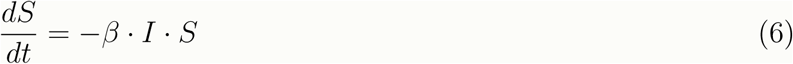

This has been solved numerically using a computerised algebra system (Maxima). If this is done with a broad skewed mask, the kinetic pattern described by the original *SIR* model is recreated with a skewed *I*. If convolved with a Kronecker delta, *N* − *S* and *R* can be shown to be an identical time shifted logistic functions such that *R*(*t*) = *N* − *S*(*t* − *λ*).

**Figure 1:**
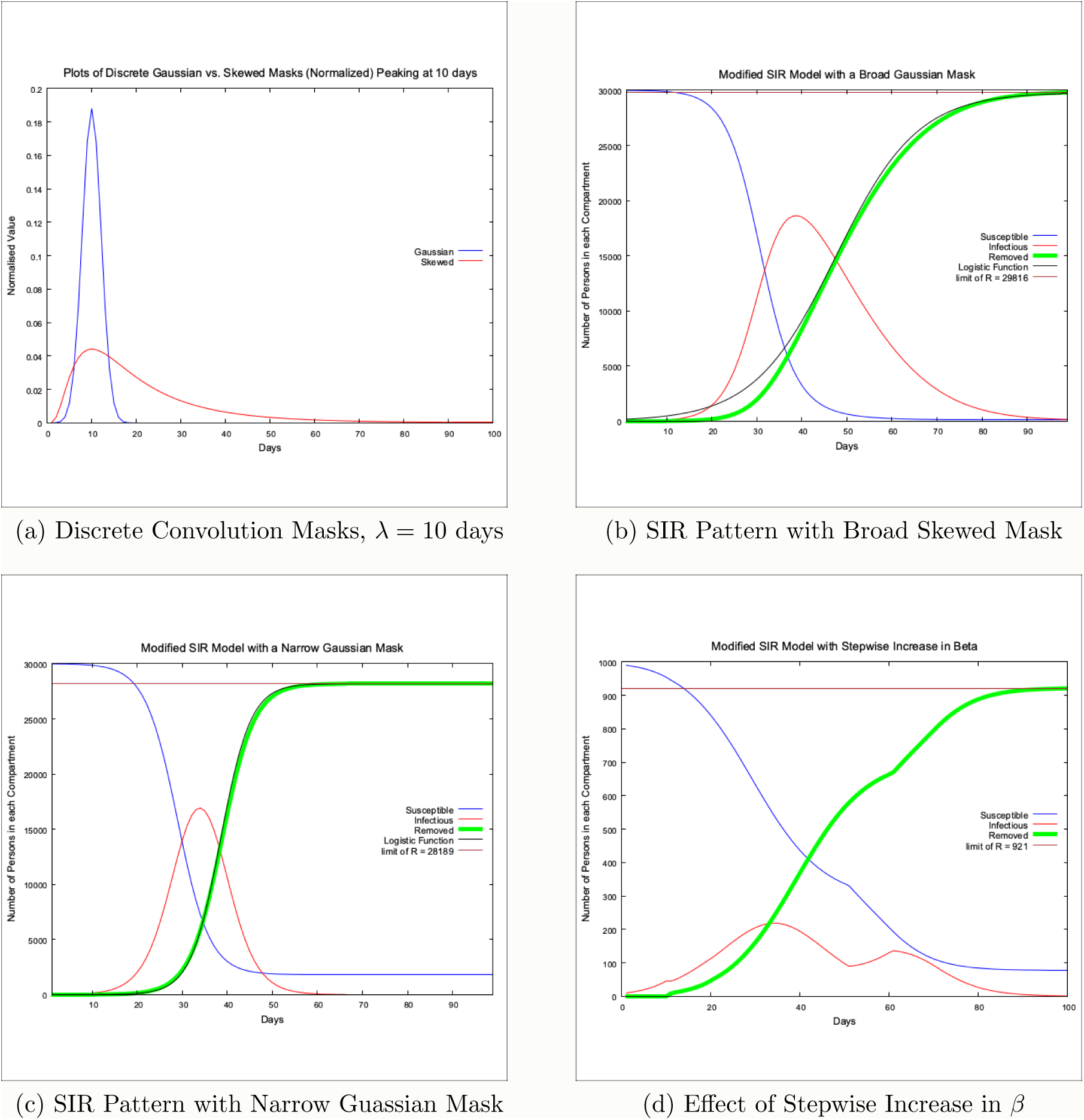
Plots of S,I and R from a numerical solution to a modified version of the tri-compartmental SIR model of Kermack and McKendrick

The original SIR model was used to model diseases that might well have a broad skewed probability distribution of latency in travelling between *S* and *R*. Many viral illnesses will have a more predictable *λ*. We should in the first instance attempt to model R with a simple logistic function.

It should be noted that, although logistic in shape, the model does not exhaust all of the susceptible population over time. In this way it differs dynamically from Verhulst’s original concept.[5] The limiting value of cumulative mortality is therefore not fixed, increasing with increasing *β* and *λ* though must be less than the initial susceptible population. This will allow for further smaller peaks of infection should *β* subsequently increase while *I* is not zero.

## 3 Modelling The Covid-19 Pandemic using a Logistic Curve

The development of suitable serological and PCR tests and the rolling out of testing has understandably trailed the rapid evolution of the Covid-19 pandemic.[6][7] Morbidity data is bound to be influenced by testing bias. Mortality data is likely to be more reliable because the probability of testing will be highest in those recently deceased following the exhibition of symptoms of the illness. Mortality *D*(*t*) to be proportional to *R*(*t*), related by an as yet unknown case fatality probability. We should, therefore, study mortality data to inform our pandemic modelling. Of course we need to be aware that mortality data will lag behind morbidity data.

Cumulative mortality is an increasing monotone. The daily geometric increase in mortality *D* is the growth factor G or alternatively (1 + *g*) and so *D_n_*_+1_ = *GD_n_* = (1 + *g*)*D_n_* = *D_n_* + *gD_n_*. We therefore have a window on the day to day value of *g* using the equation

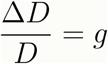

The logistic function is the solution to an ordinary differential equation 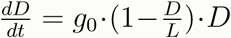 where *g*_0_ is the growth factor when *D* = 0, *L* is the limiting value of *D*. We can see that 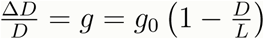 and so

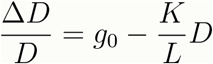

If the mortality data is modelled by a logistic we should expect a plot of 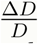 vs. *D* to be a straight line. The *D* intercept is the predicted total mortality of the pandemic. The 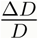 intercept is the value of *g*_0_.

We would expect the effect of social distancing to be a stepwise function modulating *g* so:

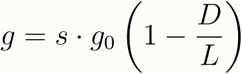

where *s* = 1 before the impact of social distancing and *s* < 1 thereafter. *s* is a stepwise function of time but as cumulative mortality vs. time is an increasing monotone the step also occurs at a particular cumulative mortality.

For any line segment in the space of 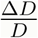 vs. *D* there is an equivalent logistic function segment in the space of *D* vs. *Date*.^1^

**Figure 2:**
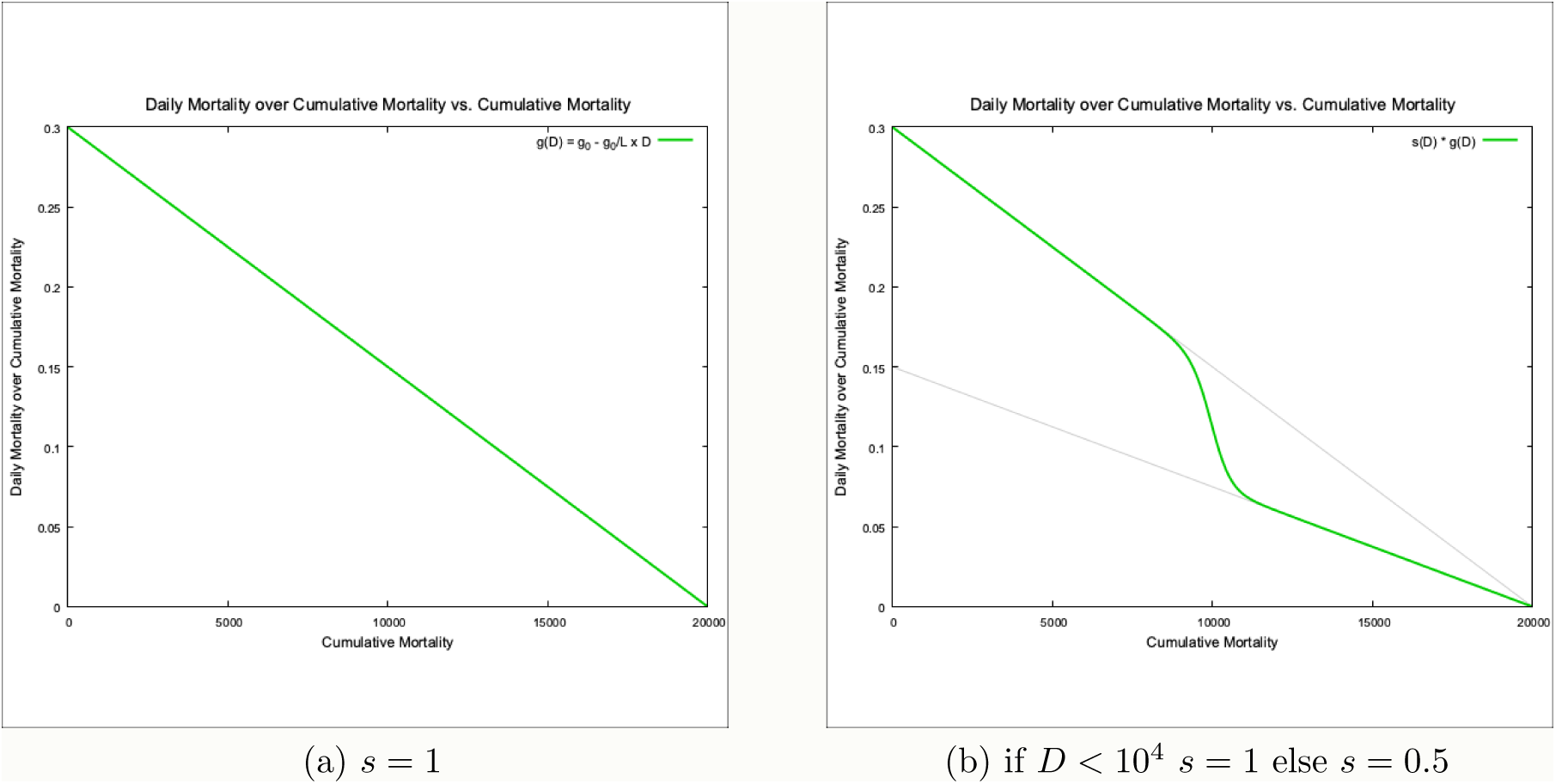
Plots of Predicted 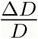 vs. *D* using a Logistic Function

## 4 Testing the Model Fit

If we look at the plots of mortality data[8] we are initially confronted by the considerable variation in the day to day reporting from each country. This is presumably related to national testing and recording procedures. For this reason the author has chosen to focus initially on the mortality data from Italy as it appears the least erratic.

### 4.1 Italy

On looking at the 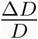 vs. *D* plot for Italy as in figure 3 we see that the data points appear to fall into two separate groups which appear highly correlated to their distinct regression lines. The segmentation into these two “phases” of the pandemic was performed algorithmically so as to avoid subjective bias, splitting the data into two at each possible location and finding a break, if present, where the 95% confidence intervals for gradient did not overlap.

**Figure 3:**
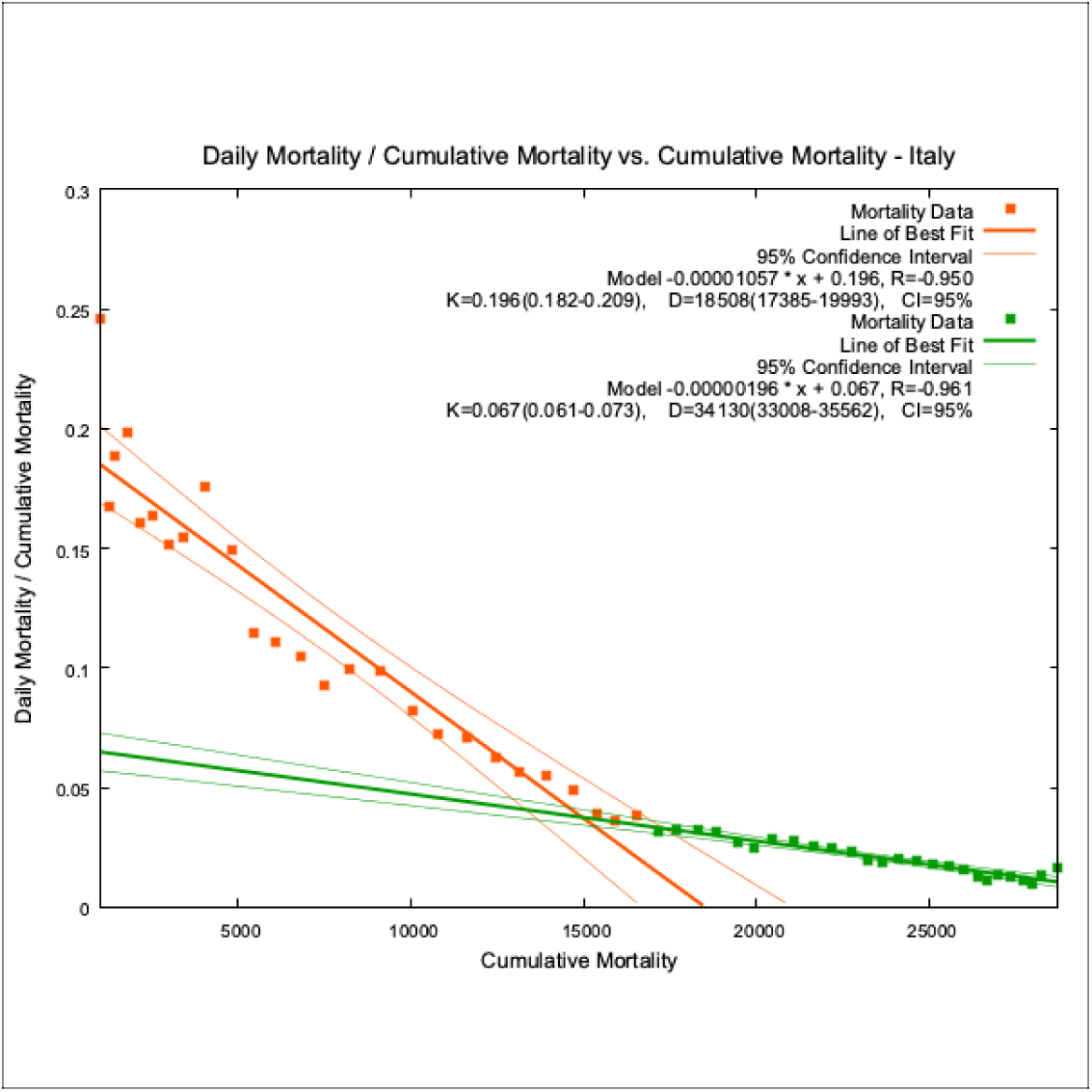
Italy, Linear Regression, Daily Mortality / Cumulative Mortality vs. Cumulative Mortality.

As previously stated, for any line segment in the space of 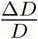 vs. *D* there is an associated logistic function segment in the space of *D* vs. *Date* (which minimises the sum of residual errors).

From figures 3, 4 and 5 we can see that the cumulative mortality data for Italy is modelled by two logistic functions spliced together in a piecewise fashion:

**Figure 4:**
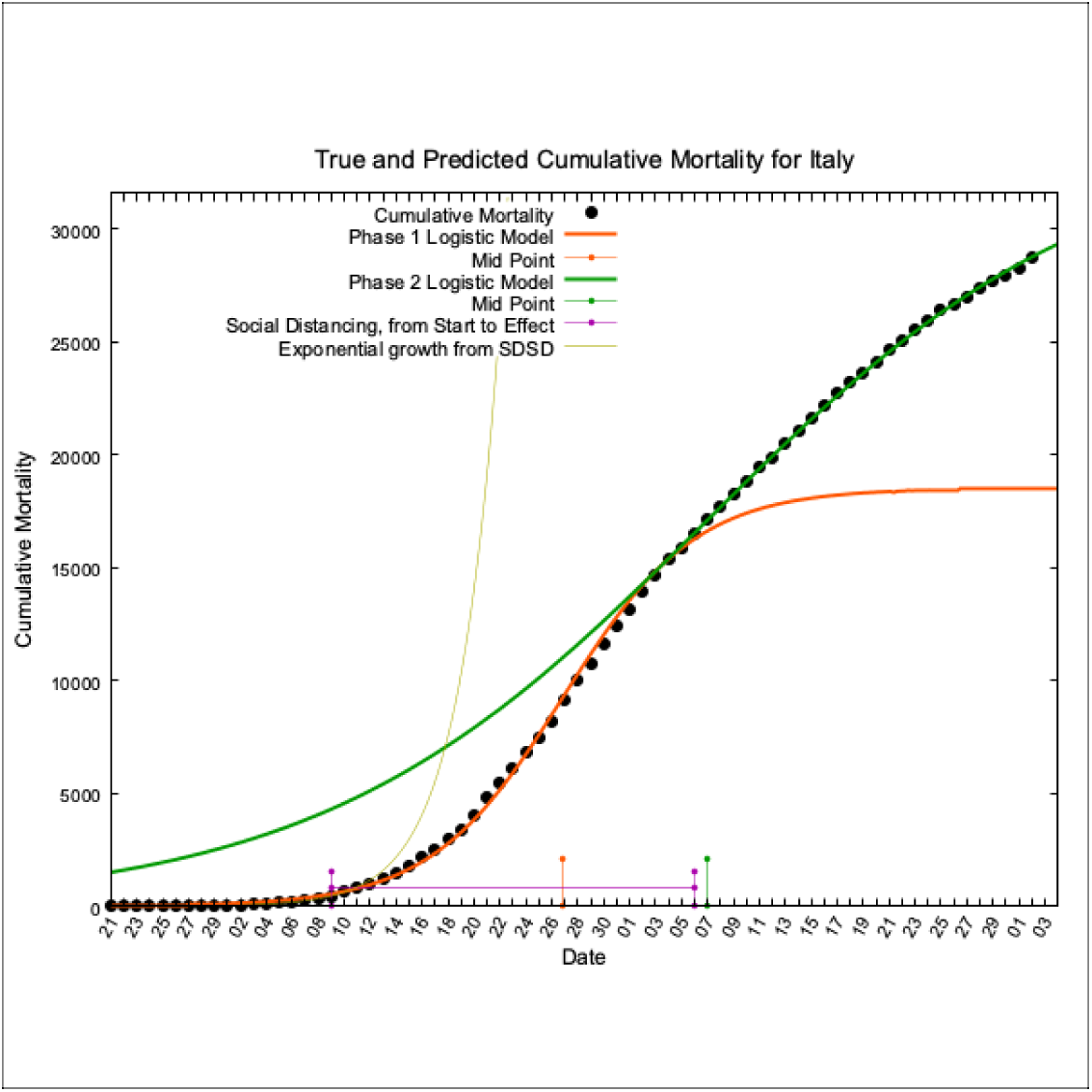
Italy, True and Modelled Cumulative Mortality vs. Date.

**Figure 5:**
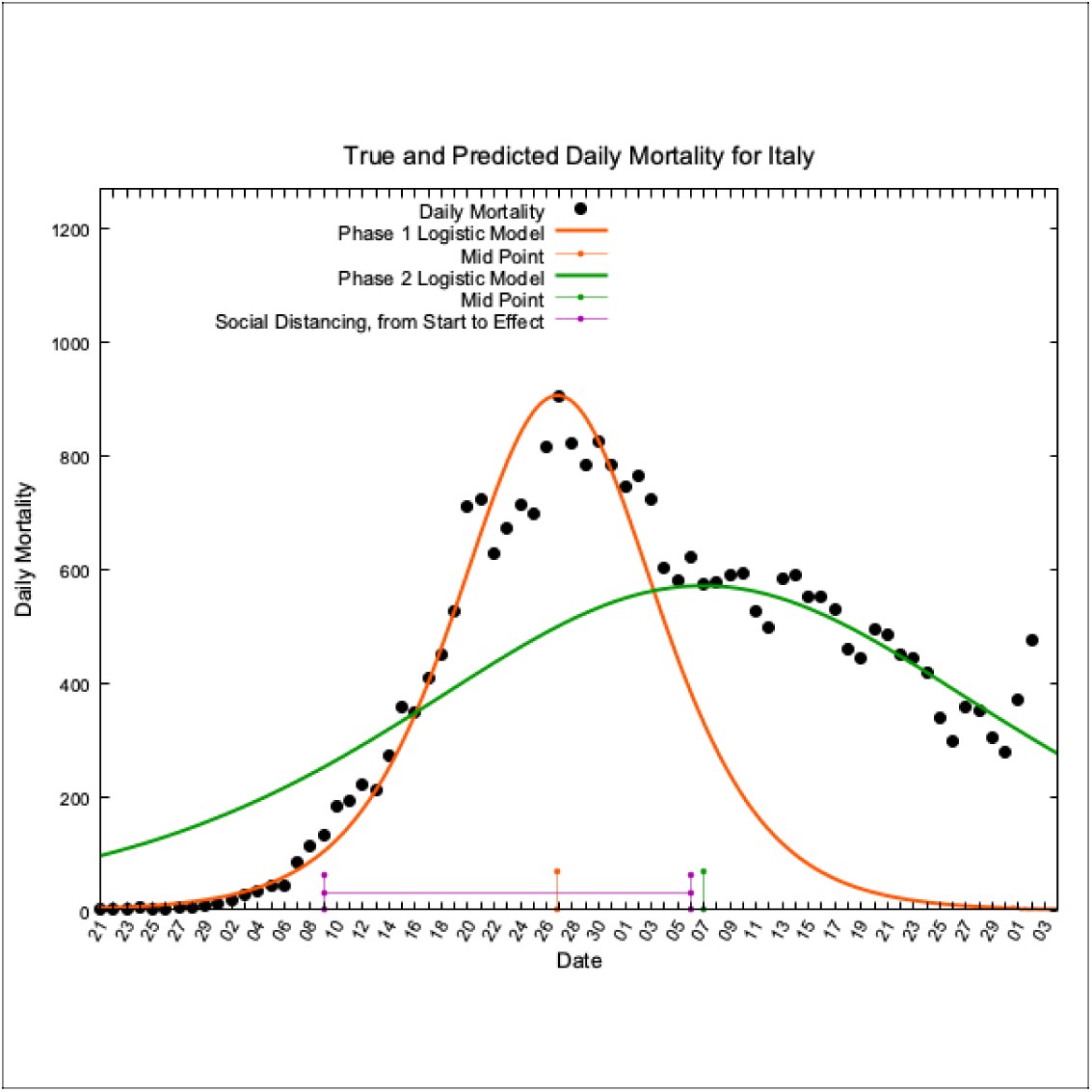
Italy, True and Modelled Daily Mortality vs. Date.

The date of peak mortality for each logistic is marked on the date axis in orange (the “natural peak”) and green (the modified peak).

An exponential growth curve is attached to the data point on the day of implementation of “lockdown” with a growth factor based on the gradient of the regression line of the preceding week’s log mortality data. It is labelled Exponential Growth from Social Distancing Start Date (SDSD). It is evident that in the period encompassing social distancing implementation, mortality growth was not exponential but rather a logistic function, which unperturbed by any other factor, would naturally flatten (and daily mortality reach its peak).

The date of the start of social distancing through “lockdown” has been marked on the plot with a purple T bar between it and the date of the natural juncture of the two logistic curves. This marks the point at which social distancing appears to have produced an abrupt stepwise reduction in *g*. For Italy this date occurs 25 days after the start of social distancing. We must assume that in Italy the average duration of time from contracting Covid-19 to death for those unfortunate enough to succumb is around 3.5 weeks.

### 4.2 Plots for Italy, Spain, France and The UK

The plots for Spain, Italy, France and the UK follow for side by side comparison:

The overall pattern of a “natural” peak followed by the remains of a modified peak is present in the data from both Italy and Spain. In the UK this pattern is just about discernible though the data has much more day to day variability. France appears to have a single phase to its pandemic with daily mortality climbing and descending a sharper “natural” peak.

**Figure 6:**
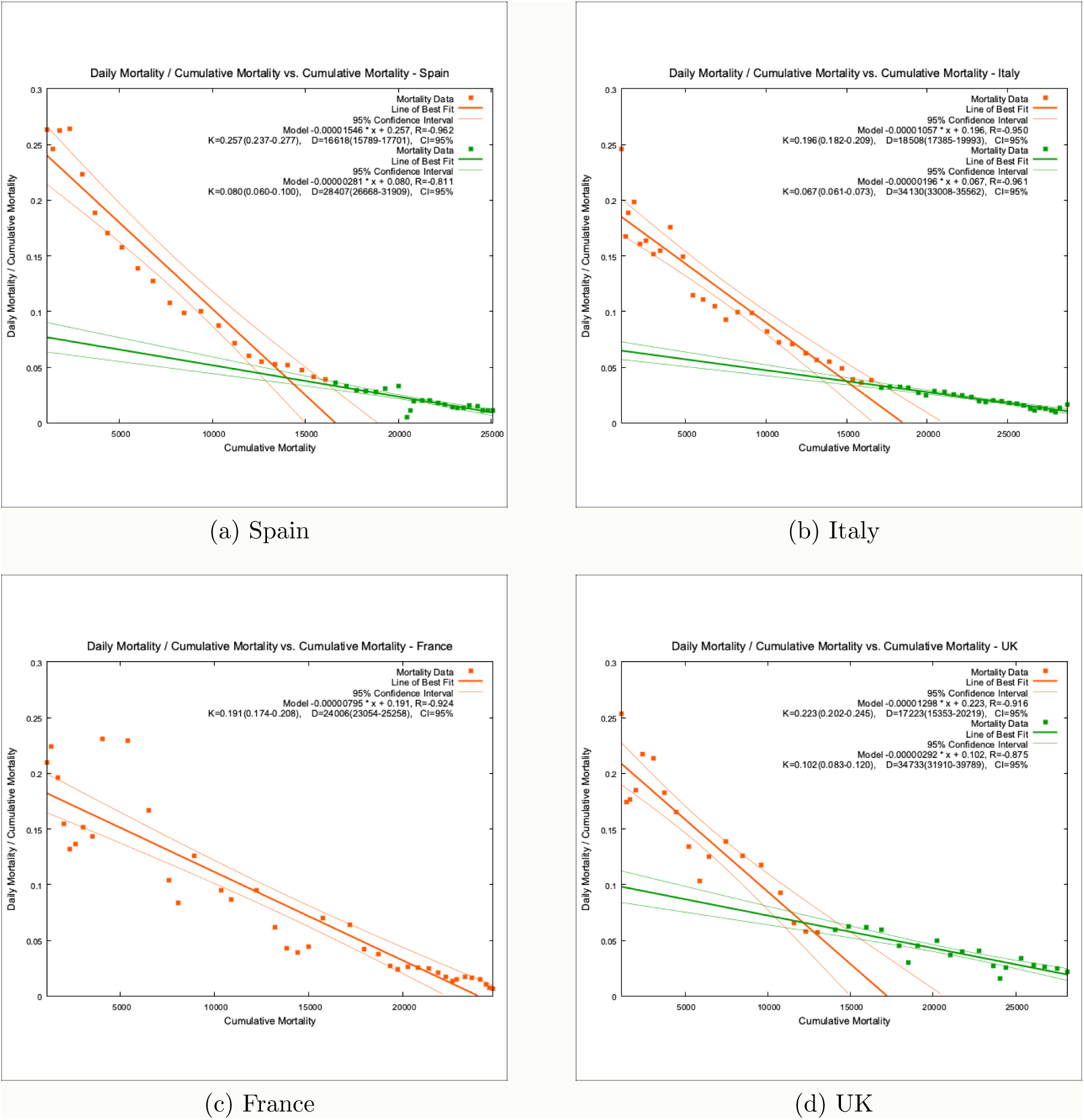
Plots of 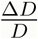 vs. *D* and their Regression Lines

**Figure 7:**
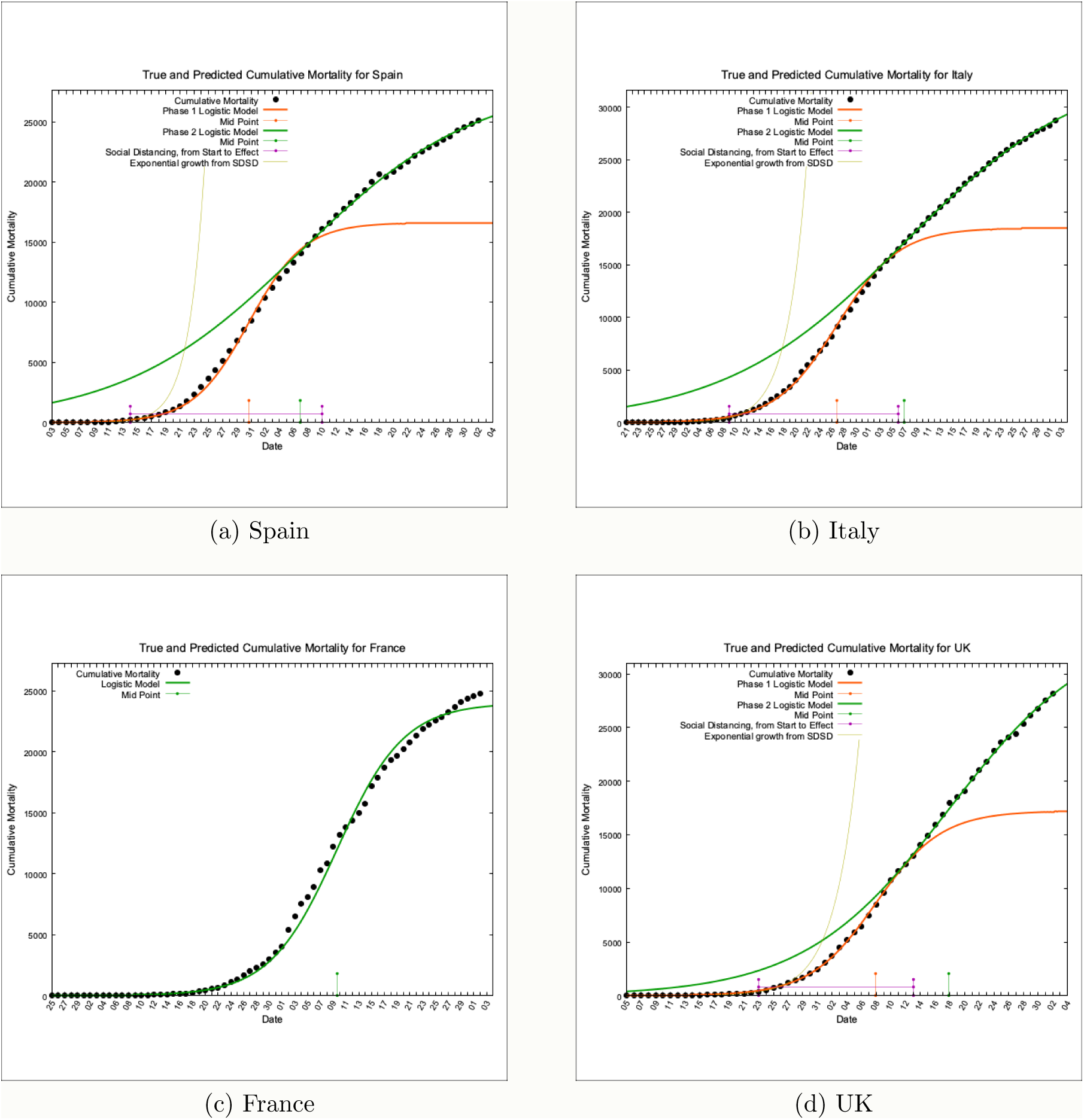
Cumulative Mortality, Actual and Predicted

**Figure 8:**
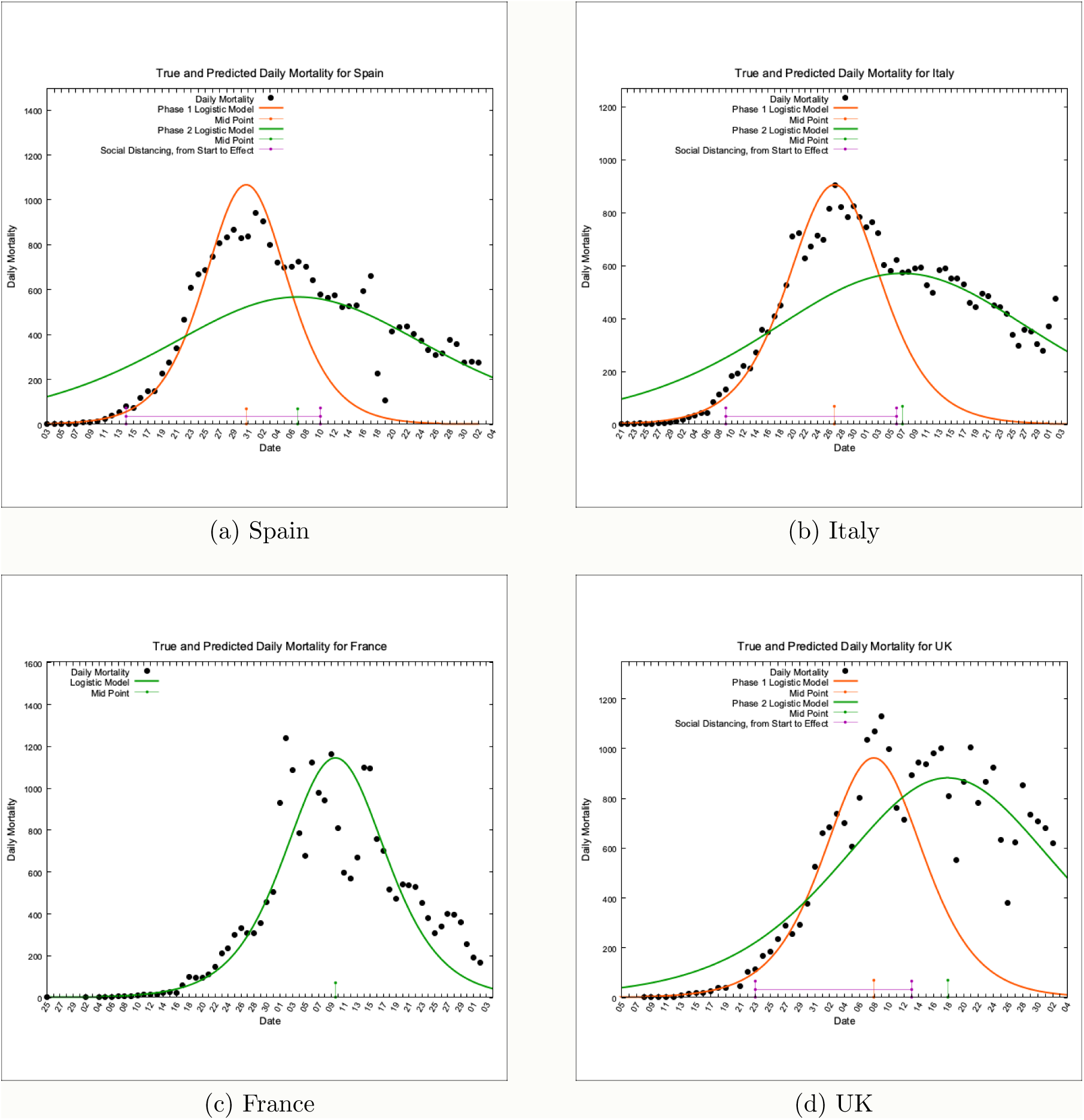
Daily Mortality, Actual and Predicted

## 5 Model Predictions

The data for Italy, Spain and the UK appear biphasic. It is not possible to discern a clearly biphasic pattern in the mortality data from France. The following tables contain the model predictions compared with true values.

**Table 1:**
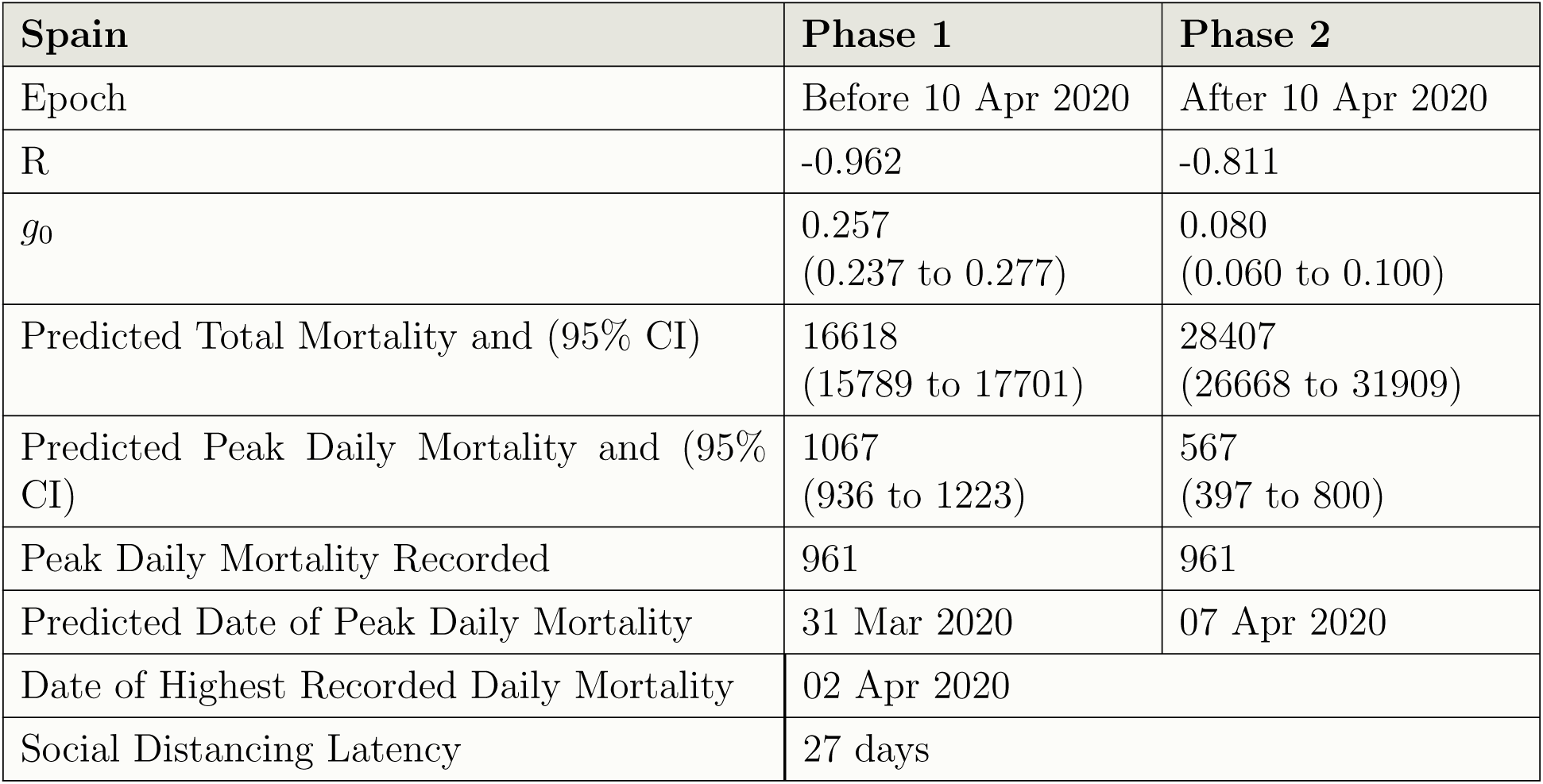
Details for Spain

**Table 2:** Details for Italy

| Italy | Phase 1 | Phase 2 |
| --- | --- | --- |
| Epoch | Before 06 Apr 2020 | After 06 Apr 2020 |
| R | -0.950 | -0.961 |
| $g_0$ | 0.196<br>(0.182 to 0.209) | 0.067<br>(0.061 to 0.073) |
| Predicted Total Mortality and (95% CI) | 18508<br>(17385 to 19993) | 34130<br>(33008 to 35562) |
| Predicted Peak Daily Mortality and (95% CI) | 905<br>(791 to 1046) | 570<br>(505 to 645) |
| Peak Daily Mortality Recorded | 919 | 919 |
| Predicted Date of Peak Daily Mortality | 27 Mar 2020 | 07 Apr 2020 |
| Date of Highest Recorded Daily Mortality | 27 Mar 2020 |  |
| Social Distancing Latency | 28 days |  |

**Table 3:**
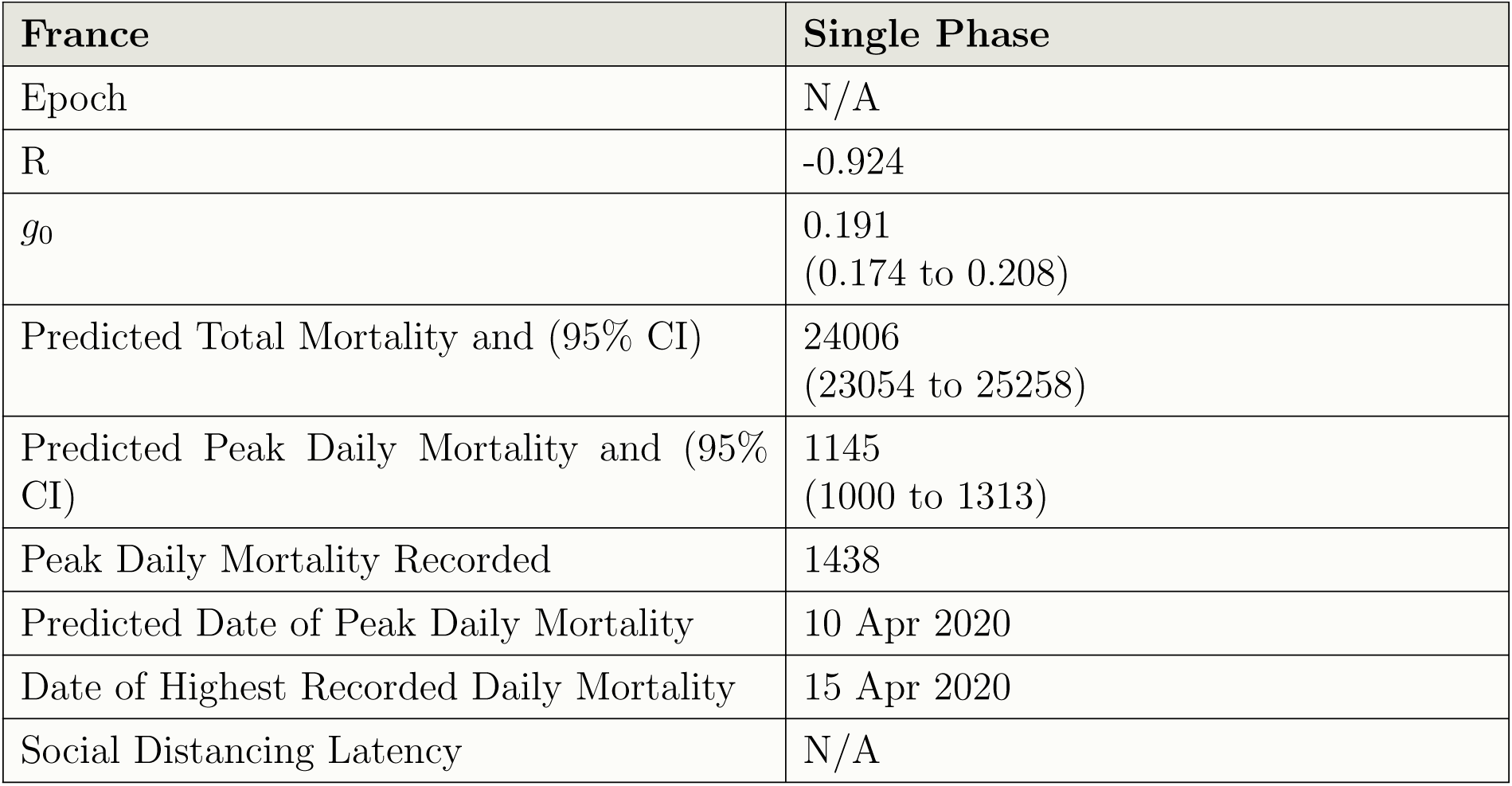
Details for France

**Table 4:** Details for UK

| UK | Phase 1 | Phase 2 |
| --- | --- | --- |
| Epoch | Before 13 Apr 2020 | After 13 Apr 2020 |
| R | -0.916 | -0.875 |
| $g_0$ | 0.223<br>(0.202 to 0.245) | 0.102<br>(0.083 to 0.120) |
| Predicted Total Mortality and (95% CI) | 17223<br>(15353 to 20219) | 34733<br>(31910 to 39789) |
| Predicted Peak Daily Mortality and (95% CI) | 962<br>(773 to 1240) | 882<br>(665 to 1191) |
| Peak Daily Mortality Recorded | 1172 | 1172 |
| Predicted Date of Peak Daily Mortality | 08 Apr 2020 | 18 Apr 2020 |
| Date of Highest Recorded Daily Mortality | 21 Apr 2020 |  |
| Social Distancing Latency | 21 days |  |

## 6 Discussion

Although we might be pleased with the fit of the model, the conclusions that inevitably follow if it correctly describes the pandemic in these countries, are both surprising and disquieting.

The principle stated aim of social distancing is to reduce peak daily mortality, allow health services to cope with those seriously affected by the disease and so save lives.[9] It is clear, however, that social distancing has not modified the attainment of the “natural” peak in daily mortality predicted by the phase 1 logistic function (the only logistic function for France) in these 4 countries. It has neither reduced the predicted peak in daily mortality nor has it altered its timing. With an average latency from the implementation of strict social distancing to an effect on mortality of 3.5 weeks, we can see that for it to have had an effect strict social distancing would have had to have been introduced at around the time of the first three deaths. This was nearly achieved in Germany with a closure of schools and colleges at 8 deaths and a blanket “lockdown” at 94 deaths. Their mortality has been much lower. We might introduce the term “Delayed Social Distancing” to distinguish it from “Timely Social Distancing”, used where social distancing occurs much later than the date of the third death from a pandemic condition.[10]

In contrast, the natural resolution of the pandemic has been delayed in Italy, Spain and the UK which now proceed along a second broader logistic. This contrasts with France’s “natural” fast up and down daily mortality peak. The predicted total mortality is higher if based on the second phase logistic (of those countries with a biphasic pattern) than that based on the first. To explore this further the ratio of the growth factors *g*_0_ for phases 1 and 2 was plotted vs. normalised predicted total mortality. 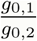 is found from the ratio of the gradient of the regression lines for pandemic phases 1 and 2 in the plots of 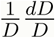 and provides a dimensionless measure of the effectiveness of strict social distancing.

**Figure 9:**
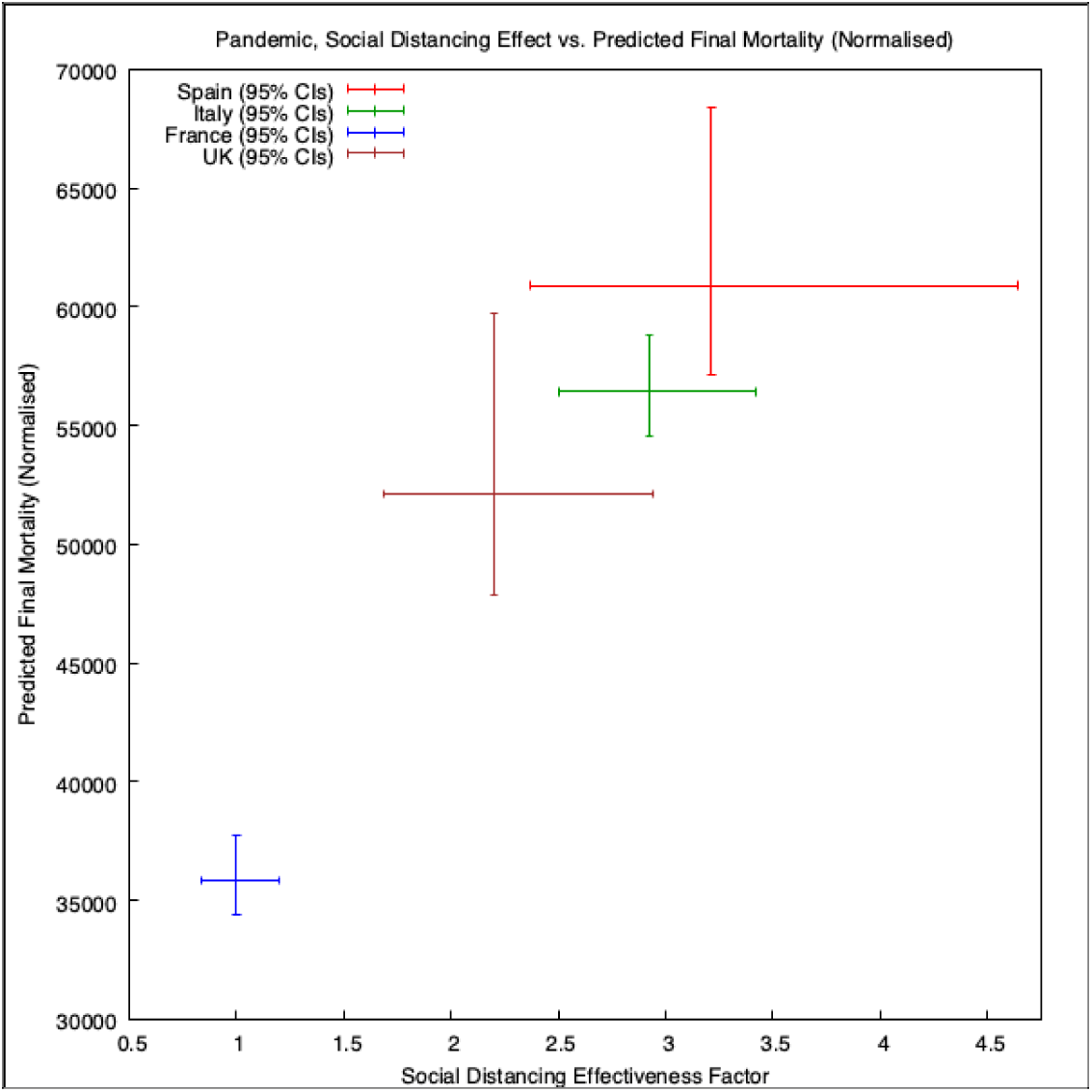
Italy, True and Modelled Cumulative Mortality vs. Date.

The old “…stable door…” adage seems surprisingly apposite. The increase in predicted total mortality with increasing social distancing effect (delayed) is quite unexpected. The pattern of mortality data might provide insight into possible underlying mechanisms. We know that certain groups are at higher risk of dying from Covid-19. Age, obesity, diabetes, gender and socio-economic status are all known factors influencing mortality in addition to the strong effect from co-morbidities. A low aerobic fitness might be a common thread amongst all these factors and is likely to be adversely affected by a “lock down”. This will be particularly true where the bulk of a person’s exercise arises from the performance of their routine activities rather than any specific training, and where there are limited opportunities at home to maintain fitness because of a lack of space or overcrowding. We already know that a modest increase in 6 minute walking distance (6MWD) has a significant impact on survival in COPD.[11]

## 7 Conclusions

If the proposed model correctly describes and predicts the evolution of the pandemic in the four countries studied then we may draw the following conclusions:

- The mortality from Covid-19 was not growing exponentially prior to the implementation of social distancing measures in any of the countries studied. The mortality data closely followed a logistic curve.
- The gradient of the logistic curve modelling the mortality data culminates in a “natural” peak. Delayed social distancing has had no impact on this “natural” peak of daily mortality which closely fits the phase 1 model peak in all the countries studied.
- To have had an effect on peak daily mortality, one of the stated principle aims of social distancing, it would have had to have been implemented at around the time of the third death from Covid-19. In countries where this was achieved, mortality has been much lower. We might use the terms timely and delayed social distancing to describe this.
- Social distancing, when implemented too late to influence the “natural” peak (as is the case in the four countries studied), appears to delay the subsequent resolution of the pandemic. The area under the curve of daily mortality is increased and so therefore is the final predicted total mortality.
- Delayed social distancing, where effective, appears to increases overall mortality. The increase in overall mortality seems directly related to the effectiveness of social distancing.
- The modified SIR model predicts that not all susceptible persons will be infected. When social distancing is relaxed we should expect a further peak in daily mortality from Covid-19. Mini-peaks of infection have been noted after the relaxation of regional social distancing used to control influenza outbreaks.[12] This is an inevitable consequence of increasing in our model. It certainly does not validate the use of delayed social distancing, nor should it influence its relaxation.

## Data Availability

Data is from the Worldometer website.

https://www.worldometers.info/coronavirus/

## Footnotes

1 There are actually an infinite number of identically shaped time shifted logistic segments in *D* vs. *Date* which would transform under this operator to a single line segment, the kernel, in 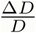 vs. *D*. The choice of time shift has been selected algorithmically to minimise RMSE

## Notes

### Competing Interest Statement

The authors have declared no competing interest.

### Funding Statement

I am not funded.

